# Extended SIR prediction of the epidemics trend of COVID-19 in Italy and compared with Hunan, China

**DOI:** 10.1101/2020.03.18.20038570

**Authors:** Jia Wangping, Han Ke, Song Yang, Cao Wenzhe, Wang Shengshu, Yang Shanshan, Wang Jianwei, Kou Fuyin, Tai Penggang, Li Jing, Liu Miao, He Yao

## Abstract

**Background:** Coronavirus Disease 2019 (COVID-19) is currently a global public health threat. Outside of China, Italy is one of the most suffering countries with the COVID-19 epidemic. It is important to predict the epidemics trend of COVID-19 epidemic in Italy to help develop public health strategies.

**Methods:** We used time-series data of COVID-19 from Jan 22,2020 to Mar 16,2020. An infectious disease dynamic extended susceptible-infected-removed (eSIR) model, which covers the effects of different intervention measures in dissimilar periods, was applied to estimate the epidemic trend in Italy. The basic reproductive number was estimated using Markov Chain Monte Carlo methods and presented using the resulting posterior mean and 95% credible interval (CI). Hunan, with similar total number of populations in Italy, was used as a comparative item.

**Results:** In the eSIR model, we estimated that the basic reproductive number for COVID-19 was respectively 4.10 (95% CI: 2.15–6.77) in Italy and 3.15(95% CI: 1.71–5.21) in Hunan. There would be totally 30 086 infected cases (95%CI:7920-81 869) under the current country blockade and the endpoint would be Apr 25 (95%CI: Mar 30 to Aug 07) in Italy. If the country blockade is imposed 5 day later, the total number of infected cases would expand the infection scale 1.50 times.

**Conclusion:** Italy’s current strict measures can efficaciously prevent the further spread of COVID-19 and should be maintained. Necessary strict public health measures be implemented as soon as possible in other European countries with a high number of COVID-19 cases. The most effective strategy needs to be confirmed in further studies.

## 1 Introduction

The Corona Virus Disease 2019 (COVID-19) started in Wuhan, China in December and quickly spread to China and many countries and regions in the world(1-3). The COVID-19 outbreak made assessment as a pandemic by the World Health Organization (WHO) on March 11. It is currently a global public health threat and more than 100 countries including Italy, Iran, the United States, South Korea, and Japan are suffering from COVID-19. Outside of China, Italy is one of the most suffering countries with the COVID-19 epidemic. As of Mar 16, the cumulative number of confirmed cases in Italy reached 27980, ranking second in the world, the total confirmed deaths, 2158, and the fatality rate, as high as 7.71%, which has become one of the highest among the major epidemic countries. However, few studies have assessed the epidemic status in Italy(4, 5).

Global public health measures are required to cope with the rapid spread of the epidemic. China has taken precise and differentiated strategies, including self-quarantine of residents in Wuhan and other areas, community-based prevention and control. These measures have played an important role in preventing and controlling the epidemic. Previous studies have shown that due to the isolation of Wuhan, the overall epidemiological progress in mainland China has been delayed by three to five days and the number of internationally transmitted cases has been reduced by nearly 80%(6). Italy detected the first two cases of imported COVID-19 on Jan 31. After that, Italy was the first country to declare a state of emergency. Since then, various measures have been implemented to control the spread of COVID-19. It is vital to evaluate the role of Italian quarantine measures for decision-making.

Mathematical modeling is helpful to predict the possibility and severity of disease outbreak and provide key information for determining the type and intensity of disease intervention. The SIR model and its modifications such as SEIR model have been widely applied to the current outbreak of COVID-19. Tang et al. estimated the infectivity of COVID-19 based on a classical susceptible-exposed-infected-removed (SEIR) epidemiological model(7). Wu et al proposed an extended SEIR model to forecast the spread of 2019-nCoV both within and outside of mainland China(3). However, these studies assumed that the exposed population were not infectious, which may be not suitable in COVID-19. Yang Z et al. predicted that China’s epidemic will peak in late February and end in late April by a combination of SEIR model and a machine-learning artificial intelligence (AI) approach(8). However, this study and the above studies did not consider the phase-adjusted preventive measures and time-varying parameters, which may affect the accuracy of predictions.

We adopted extended susceptible-infected-removed (eSIR) model(9), which covers the effects of different epidemic prevention measures in different periods and helps to achieve the following specific objectives:

AIM 1: Compare the epidemic development of COVID-19 in Italy with provinces with similar total number of populations in China.

AIM 2: Predict the epidemiological trend of COVID-19 in Italy via a modified and calibrated model.

AIM 3: Assess the effectiveness of Italian existing and assumed precautionary quarantine measures.

## 2 Methods

### 2.1 Data sources

In this study, we used the publicly available dataset of COVID-19 provided by the Johns Hopkins University(10). This dataset includes many countries’ daily count of confirmed cases, recovered cases and deaths. As time-series data, it is available from 22 January 2020. Besides, we also gathered and cross-checked data in DXY.cn(11), a website providing real-time data of the COVID-19.

These data are collected through public health authorities’ announcements and are directly reported public and unidentified patient data, so ethical approval is not required.

### 2.2 Prediction models

The reproduction number, R0, reflects the transmissibility of a virus spreading under no control, representing the average number of new infections generated by each infected person(12). The COVID-19 is likely to decline and eventually disappear if R0≤1.To estimate trends and calculate the R0, we used an extended SIR model (eSIR model) with a time-varying transmission rate(9). In brief, this model is one extension to the basic epidemiological model——susceptible-infected-removed (SIR) model. S, I and R represent the number of susceptible, infected, and recovered individuals accordingly, and N = S + I + R is the total population. The transmission rate is constant in the SIR model. It should be noted that in actual situations, the speed of transmission can be changed through many interventions, such as personal protective measures, community-level isolation and city blockade. As is shown below, the eSIR model adds transmission modifier π(t) to the SIR model, so it allows a time-varying probability of the transmission rate.

An extended SIR model with time-varying transmission rates.

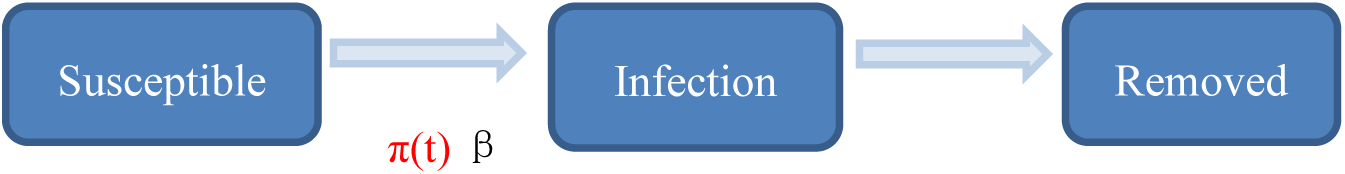

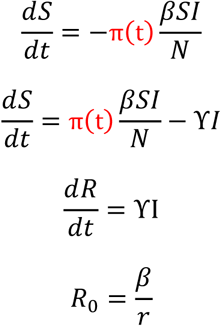

The transmission rate modifier π(t) can be specified according to actual interventions in different times and regions. According to Chinese government isolation measures and previous study, we set π(t)=0.9 if t ∈ (Jan 23, Feb 04], city blockade; π(t)=0.5 if t ∈ (Feb 4, Feb 8], enhanced quarantine; π(t)=0.1 if t >Feb 8, more enhanced quarantine in Hunan. In the opinion of Italy government isolation measures, we set π(t)=0.9 if t <Mar 4, no concrete quarantine protocols; π(t)=0.5 if t ∈ (Mar 04, Mar 09], some cities blockade and enhanced quarantine; π(t)=0.1 if t > Mar 09, country blockade and more enhanced quarantine in Italy. We also assumed that the Italy government took country blockade and intensified quarantine on Mar 05 or Mar 15.

We did prediction with an R software package—eSIR which can output the Markov Chain Monte Carlo (MCMC) estimation, inference, and prediction under the extended SIR models with time-varying transmission modifier π(t). The model can also yield the turning points of the epidemiological trend of COVID-19. The first turning point was defined as the mean predicted time when the daily proportion of infected cases becomes smaller than the previous ones. The second turning point was defined as the mean the predicted time when the daily proportion of removed cases (i.e. both recovered and dead) becomes larger than that of infected cases. Besides, an end point was defined as the time when the median proportion of current infected cases turn to zero.

We did all analyses in R (version 3.6.2).

## 3 Results

### 3.1 Epidemic development of COVID-19 in Italy compared with Hunan

Figure 1 demonstrates daily new COVID-19 cases and epidemic distribution of COVID-19 in Hunan, China and Italy. The number of new cases and confirmed cases show an exponential trend since Feb 21 in Italy from while the number of new cases turns to zero from Feb 29 in Hunan.

**Figure 1.**
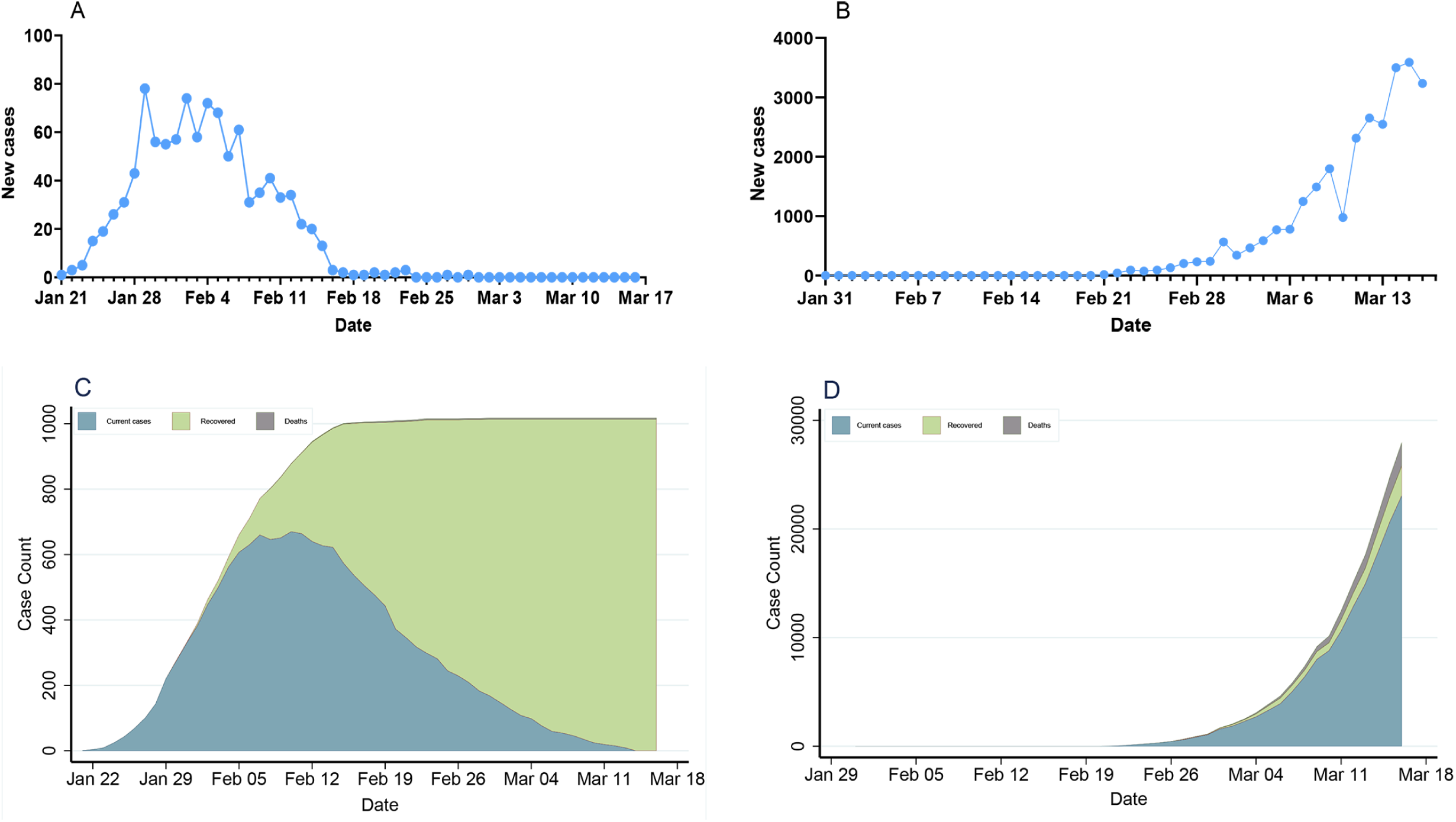
Epidemic development of COVID-19 in Hunan, China and Italy. A, B: Daily new COVID-19 cases in Hunan, China and Italy. C, D: Epidemic distribution of COVID-19 in Hunan, China and Italy.

### 3.2 Prediction of the epidemics trend of COVID-19 in Italy compared with Hunan

Table 1 summarizes the posterior mean and credible intervals of R0, incidence, and endpoint in Hunan and Italy according to the eSIR model. R0 for COVID-19 was respectively 3.15(95% CI: 1.71–5.21) in Hunan and 4.10 (95% CI: 2.15–6.77) in Italy. There would be totally 3 988 infected cases (95%CI:786-9 065) in Hunan. There would be totally 30 086 infected cases (95%CI:7 920-81 869) under the current country blockade in Italy. Figure 2 indicates an epidemiological trend of COVID-19 under existing preventions in Hunan, China and Italy. The first and second turning point in Hunan appeared on Feb 04 and Feb 09. The first and second turning point in Italy is Mar 10 and Mar 11. The predictions suggest that the endpoints of the COVID-19 epidemics in Hunan and Italy will come on Feb 23 (95%CI: Feb 19 to Mar 23) and Apr 25 (95%CI: Mar 30 to Aug 07), separately.

**Table 1.** R0, incidence and endpoint in Hunan and Italy according to the eSIR model.

| Model | R0 |  | Incidence (%) |  | Endpoint |  |
| --- | --- | --- | --- | --- | --- | --- |
|  | Mean | 95%CI | Mean | 95%CI | Mean | 95%CI |
| Hunan | 3.15 | 1.71-5.21 | 0.0066 | 0.0013-0.0150 | 2020/2/23 | 2020/2/19-2020/3/23 |
| Italy | 4.10 | 2.15-6.77 | 0.0498 | 0.0131-0.1355 | 2020/4/25 | 2020/3/30-2020/8/07 |

Inf: The endpoint appears more than 200 days after $t_0$ .

**Figure 2.**
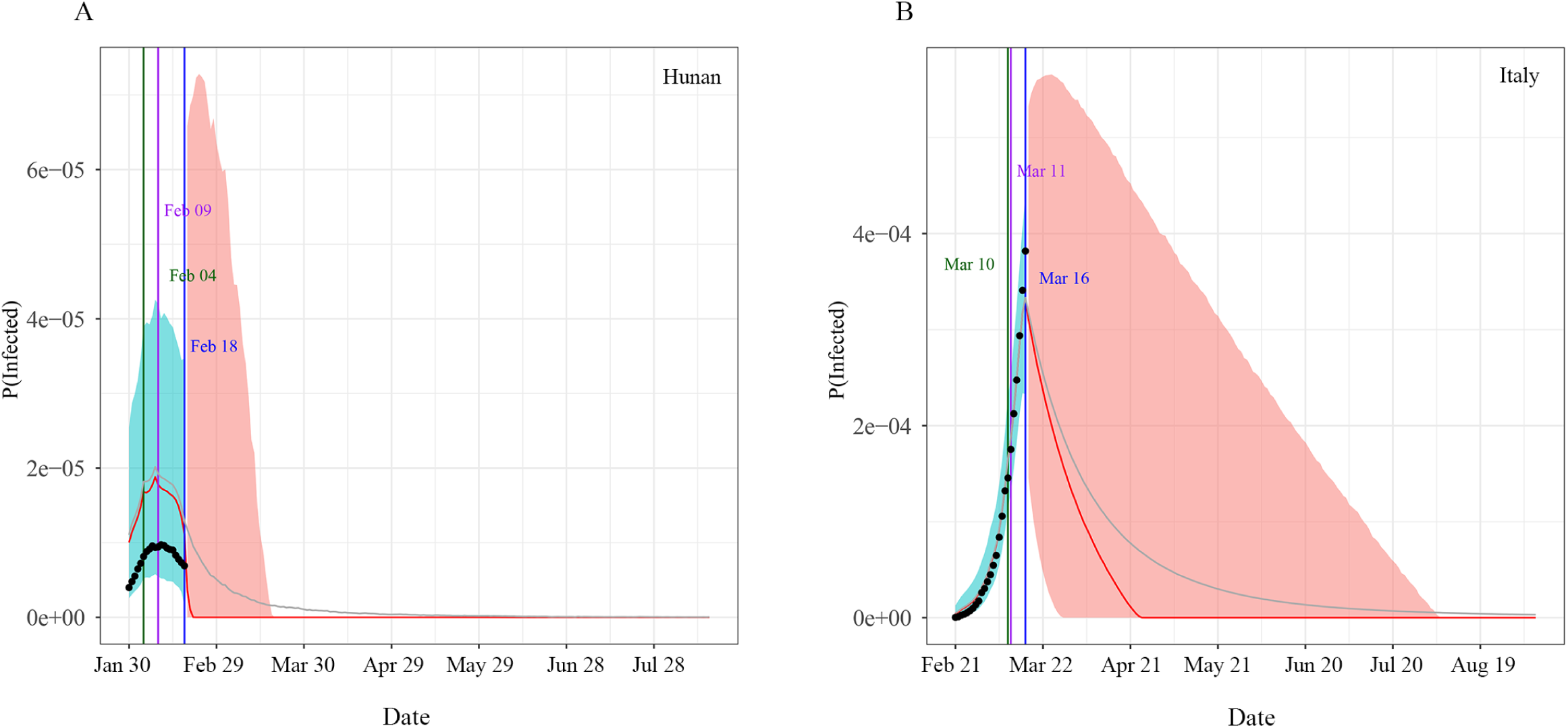
Epidemiological trend of COVID-19 under existing preventions in Hunan, China and Italy according to eSIR model. A: Hunan, China; B: Italy. The black dots left to the blue vertical line denote the observed proportions of the infected and removed compartments on the last date of available observations or before. The blue vertical marks is the current time up to which we have observed data (t_0_). The green and purple vertical lines denote the first and second turning points, respectively. The cyan and salmon color area denotes the 95% credible interval of the predicted proportions of current infected cases before and after t_0_, respectively. The gray and red curves are the posterior mean and median curves. Epidemiological trend of COVID-19 under existing preventions of Hunan, China and Italy in eSIR model

### 3.3 Prediction of the epidemics trend of COVID-19 in Italy assumed precautionary quarantine measures

Table 2 summarizes the posterior mean and credible intervals of R0, incidence, and endpoint according to different beginning of country blockade in Italy. The mean number of COVID-19 infected cases in Italy would be 10636(95%CI: 2357-23326) and the endpoint would arrive on Mar 16 (95%CI: Mar 09 to Apr 22) if the beginning time of country blockade were Mar 05. Furthermore, that would be 44993(95%CI: 16676-96759) and Apr 27 (95%CI: Apr 01 to Jul 27) if the beginning of country blockade were Mar 15.

**Table 2.** R0, incidence and endpoint in Italy according to different beginning of country blockade.

| Begin time of<br>country blockade | R0 |  | Incidence (%) |  | Endpoint |  |
| --- | --- | --- | --- | --- | --- | --- |
|  | Mean | 95%CI | Mean | 95%CI | Mean | 95%CI |
| Mar 05 | 3.26 | 1.83-5.34 | 0.0176 | 0.0039-0.0386 | 2020/3/16 | 2020/3/9-2020/4/22 |
| Mar 15 | 4.24 | 2.44-6.61 | 0.0745 | 0.0028-0.1601 | 2020/4/27 | 2020/4/1-2020/7/26 |

## 4 Discussion

This impact of the COVID-19 response (overall quarantine regulations, social distancing, and isolation of infections) in China is encouraging for the many other countries(13).We compared the situation in Hunan, China, which has the similar population as Italy to predict. The spread of COVID-19 in Hunan Province appeared relatively early and has now entered a phase of no inflections, which helps to observe the entire course of the epidemic. Besides, due to the similarity of population size and geographical location adjacent to Hubei, Hunan’s public health measures can provide useful guidance for Italy in preventing the further spread of COVID-19.

In our study, the eSIR model with R software package was used to evaluate the impact of intervention measures on Italian COVID-19 epidemic. In previous studies, estimation of the epidemic of an infectious disease is often performed using constant parameter(14-17). The advantage of eSIR model is that it combines time-varying isolation measures and expands SIR model to adapt to the time-varying transmission rate in the population. Lili Wang et al. found that COVID-19 outside Hubei in China has been so far much less severe.(9) But they did not perform each province’s analyses. The first and second points in our study are respectively Feb 04 and Feb 09,which are the same as these outside Hubei in China. Furthermore, the actual number of infected cases (1018) is included in the predicted number of infected cases (786-9065) and the endpoint(Feb 29) is included in the predicted endpoint(Feb 19 to Mar 23) in our study, which also reflects stability and accuracy of the eSIR model. Combining the above data and methods, these findings show that eSIR model is more suitable for predicting the epidemic trend of COVID-19.

Our results showed that R0 was estimated to be respectively 3.12 (95% CI, 1.83-3.01) and 3.74 (95% CI, 2.09-6.02) in SIR model and eSIR model in Italy; 2.97 (95% CI, 1.80-5.01) and 3.15 (95% CI, 1.71-5.21) in SIR model and eSIR model in Hunan. Li Qun et al estimated R0 to be 2.2 (95% CI, 2.09-6.02) among the first 425 patients in Wuhan, China(18). Other studies estimated R0 to be 1.4-2.5 (19), 2.68(95% CI 2.47∼2.68) (3), 3.6-3.8(20), 6.47(95% CI 5.71-7.23) (7). Ying Liu et al found that the estimated mean R0 for COVID-19 is around 3.28, with a median of 2.79 and IQR of 1.16 by reviewing R0 of COVID-19 in 12 studies. (21) We estimated R0 to be 3.15(95% CI 1.71-5.21) in Hunan, which is in agreement with these findings. But R0 in Italy was estimated to be 4.10 (95% CI: 2.15–6.77), which is larger than that in Hunan. This needs to be confirmed by further studies.

This study showed that COVID-19 spread rapidly throughout Italy after Feb 21. Possible reasons for such rapid growth of infections include:(1) more timely caution and preventative measures were not taken, (2) The number of infections during Jan 31-Feb 20 could be under-reported due to underdiagnosis, given subclinical or asymptomatic cases. The incubation period for COVID-19 is thought to be within 14 days following exposure, with most cases occurring approximately four to five days after exposure(18, 22, 23). So it seems impossible to maintain totally two or three cases during Jan 31-Feb 20 in Italy. In addition, the rapid increase in the number of infections after Feb 21 might reflect a belated realization of the spread of COVID-19.

Previous studies have shown that more rigorous government control policies were associated with a slower increase in the infected population(6, 16, 24-27). Our report also showed that rigorous measures can effectively prevent the further spread of COVID-19 in Hunan. Based on our model, Italy should still maintain all levels of quarantines like China by Apr 25(Mar 30-Aug 07). Furthermore, Tianyi Qiu et al found that delaying the lockdown by 1-6 days in Wuhan would expand the infection scale 1.23-4.94 times and the epidemic would be finally out of control if lockdown imposed 7 days later(17). Our study also shows that taking government control earlier can decrease the number of infected cases. In addition, from China’s experience, various control measures, including the early detection and isolation of individuals with symptoms, traffic restrictions, medical tracking, and entry or exit screening, can well prevent the further spread of COVID-19. These measures are in line with the latest recommendations by the World Health Organization and a previous study in Spain(28). But the most effective strategy still needs to be confirmed by further studies. Consequently, it is better and necessary to apply strict public health measures in other European countries with a high number of COVID-19 cases.

Our study has some limitations. Firstly, it is based on the assumption that rigorous measures like China have been taken in Italy, although this study uses the new model to obtain dynamic results, which is instructive for the prevention and control of the epidemic in Italy. Secondly, the suspected cases and the daily number of hospitalized cases are not available, so they are not considered in the eSIR model. Thirdly, some unforeseeable factors may affect these estimated data in our study such as super-spreaders exist.

In conclusion, the current study is the first to provide a prediction for epidemic trend after strict prevention and control measures were implemented in Italy. Our study suggests that rigorous measures like China should still be maintained in Italy by Apr 25(Mar 30-Aug 07) to prevent further spread of COVID-19.

## Data Availability

We used the publicly available dataset of COVID-19 provided by the Johns Hopkins University

https://github.com/CSSEGISandData/COVID-19

## 7 Author Contributions

JW, HK, LM and HY contributed to the study design. JW, HK, and LM contributed to the writing of the manuscript. JW, HK, SY, CW contributed to the data analysis. SY, CW and WJ contributed to the data compilation. WJ, YS, WS and HY contributed to critical review.TP, KF and LJ contributed to the literature search. YS, WJ, and KF contributed to the design of tables and figures.

## 8 Acknowledgments

We greatly appreciated the technical assistance provided by Zhenxing Cheng; Institute of Blue and Green Development, Shandong University, Weihai, 264209, PR China.

## Reference

1. Benvenuto D, Giovanetti M, Salemi M, Prosperi M, De Flora C, Junior Alcantara LC, et al. The global spread of 2019-nCoV: a molecular evolutionary analysis. Pathog Glob Health (2020):1–4. doi: 10.1080/20477724.2020.1725339. PubMed PMID: 32048560.

2. Liao X, Wang B, Kang Y. Novel coronavirus infection during the 2019-2020 epidemic: preparing intensive care units-the experience in Sichuan Province, China. Intensive Care Med (2020). doi: 10.1007/s00134-020-05954-2. PubMed PMID: 32025779.

3. Wu JT, Leung K, Leung GM. Nowcasting and forecasting the potential domestic and international spread of the 2019-nCoV outbreak originating in Wuhan, China: a modelling study. Lancet (2020). doi: 10.1016/S0140-6736(20)30260-9. PubMed PMID: 32014114.

4. Giovanetti M, Benvenuto D, Angeletti S, Ciccozzi M. The first two cases of 2019-nCoV in Italy: Where they come from? J Med Virol (2020):10.1002/jmv.25699. doi: 10.1002/jmv.25699. PubMed PMID: 32022275.

5. Porcheddu R, Serra C, Kelvin D, Kelvin N, Rubino S. Similarity in Case Fatality Rates (CFR) of COVID-19/SARS-COV-2 in Italy and China. J Infect Dev Ctries (2020) 14(2):125–8. doi: 10.3855/jidc.12600. PubMed PMID: 32146445.

6. Chinazzi M, Davis JT, Ajelli M, Gioannini C, Litvinova M, Merler S, et al. The effect of travel restrictions on the spread of the 2019 novel coronavirus (COVID-19) outbreak. Science (New York, NY) (2020).

7. Tang B, Wang X, Li Q, Bragazzi NL, Tang S, Xiao Y, et al. Estimation of the Transmission Risk of the 2019-nCoV and Its Implication for Public Health Interventions. J Clin Med (2020) 9(2). doi: 10.3390/jcm9020462. PubMed PMID: 32046137.

8. Yang Z, Zeng Z, Wang K, Wong S-S, Liang W, Zanin M, et al. Modified SEIR and AI prediction of the epidemics trend of COVID-19 in China under public health interventions. 2020 (2020).

9. Song PX, Wang L, Zhou Y, He J, Zhu B, Wang F, et al. An epidemiological forecast model and software assessing interventions on COVID-19 epidemic in China. medRxiv (2020):2020.02.29.20029421. doi: 10.1101/2020.02.29.20029421.

10. Dong E, Du H, Gardner L. An interactive web-based dashboard to track COVID-19 in real time. The Lancet Infectious diseases (2020). Epub 2020/02/23. doi: 10.1016/s1473-3099(20)30120-1. PubMed PMID: 32087114.

11. Sun K, Chen J, Viboud C. Early epidemiological analysis of the coronavirus disease 2019 outbreak based on crowdsourced data: a population-level observational study. The Lancet Digital Health (2020). doi: 10.1016/s2589-7500(20)30026-1.

12. Imai, N. et al. Report 3: Transmissibility of 2019-nCoV.https://www.imperial.ac.uk/mrc-global-infectious-disease-analysis/news–wuhan-coronavirus/ (2020).

13. Anderson RM, Heesterbeek H, Klinkenberg D, Hollingsworth TD. How will country-based mitigation measures influence the course of the COVID-19 epidemic? Lancet (2020):S0140-6736(20)30567-5. doi: 10.1016/S0140-6736(20)30567-5. PubMed PMID: 32164834.

14. Zhao S, Lin Q, Ran J, Musa SS, Yang G, Wang W, et al. Preliminary estimation of the basic reproduction number of novel coronavirus (2019-nCoV) in China, from 2019 to 2020: A data-driven analysis in the early phase of the outbreak. International Journal of Infectious Diseases (2020) 92:214–7. doi: https://doi.org/10.1016/j.ijid.2020.01.050.

15. Roosa K, Lee Y, Luo R, Kirpich A, Rothenberg R, Hyman JM, et al. Real-time forecasts of the COVID-19 epidemic in China from February 5th to February 24th, 2020. Infectious Disease Modelling (2020) 5:256-63. Epub 2020/02/29. doi: 10.1016/j.idm.2020.02.002. PubMed PMID: 32110742; PubMed Central PMCID: PMCPMC7033348.

16. Fang Y, Nie Y, Penny M. Transmission dynamics of the COVID-19 outbreak and effectiveness of government interventions: A data-driven analysis. J Med Virol (2020). Epub 2020/03/07. doi: 10.1002/jmv.25750. PubMed PMID: 32141624.

17. Wan H, Cui J-a, Yang G-J. Risk estimation and prediction by modeling the transmission of the novel coronavirus (COVID-19) in mainland China excluding Hubei province. medRxiv (2020):2020.03.01.20029629. doi: 10.1101/2020.03.01.20029629.

18. Li Q, Guan X, Wu P, Wang X, Zhou L, Tong Y, et al. Early Transmission Dynamics in Wuhan, China, of Novel Coronavirus-Infected Pneumonia. The New England journal of medicine (2020). doi: 10.1056/NEJMoa2001316. PubMed PMID: MEDLINE:31995857.

19. Mahase E. China coronavirus: what do we know so far? BMJ (Clinical research ed) (2020) 368:m308. Epub 2020/01/26. doi: 10.1136/bmj.m308. PubMed PMID: 31980434.

20. Read, J. M., Bridgen, J. R. E., Cummings, D. A. T., Ho, A. & Jewell, C. P. Novel coronavirus 2019-nCoV: early estimation of epidemiological parameters and epidemic predictions. medrxiv. https://www.medrxiv.org/content/10.1101/2020.01.23.20018549v1.full.pdf (2020).

21. Liu Y, Gayle AA, Wilder-Smith A, Rocklöv J. The reproductive number of COVID-19 is higher compared to SARS coronavirus. J Travel Med (2020). doi: 10.1093/jtm/taaa021. PubMed PMID: 32052846.

22. Guan WJ, Ni ZY, Hu Y, Liang WH, Ou CQ, He JX, et al. Clinical Characteristics of Coronavirus Disease 2019 in China. The New England journal of medicine (2020). Epub 2020/02/29. doi: 10.1056/NEJMoa2002032. PubMed PMID: 32109013.

23. Chan JF, Yuan S, Kok KH, To KK, Chu H, Yang J, et al. A familial cluster of pneumonia associated with the 2019 novel coronavirus indicating person-to-person transmission: a study of a family cluster. Lancet (2020) 395(10223):514-23. Epub 2020/01/28. doi: 10.1016/s0140-6736(20)30154-9. PubMed PMID: 31986261.

24. Wang H, Wang Z, Dong Y, Chang R, Xu C, Yu X, et al. Phase-adjusted estimation of the number of Coronavirus Disease 2019 cases in Wuhan, China. Cell Discovery (2020) 6(1):10. doi: 10.1038/s41421-020-0148-0.

25. Kraemer MUG, Yang C-H, Gutierrez B, Wu C-H, Klein B, Pigott DM, et al. The effect of human mobility and control measures on the COVID-19 epidemic in China. medRxiv (2020):2020.03.02.20026708. doi: 10.1101/2020.03.02.20026708.

26. Nishiura H, Kobayashi T, Miyama T, Suzuki A, Jung S, Hayashi K, et al. Estimation of the asymptomatic ratio of novel coronavirus infections (COVID-19). medRxiv (2020):2020.02.03.20020248. doi: 10.1101/2020.02.03.20020248.

27. Tian H, Liu Y, Li Y, Wu C-H, Chen B, Kraemer MUG, et al. The impact of transmission control measures during the first 50 days of the COVID-19 epidemic in China. medRxiv (2020):2020.01.30.20019844. doi: 10.1101/2020.01.30.20019844.

28. Aleta A, Moreno Y. Evaluation of the potential incidence of COVID-19 and effectiveness of contention measures in Spain: a data-driven approach. medRxiv (2020):2020.03.01.20029801. doi: 10.1101/2020.03.01.20029801.

